# Subtle oromotor signs in early Parkinson’s disease before clinical manifestations of dysphagia

**DOI:** 10.1101/2021.02.19.21251538

**Authors:** Sunita Gudwani, Prabhakar Upadhyay, Kamlesh Sharma, Rajesh Sagar, S. Senthil Kumaran, R.K. Dhamija

## Abstract

**Background:** Swallowing is complex phenomena comprising oral (preparatory and pre-swallow positioning), oropharyngeal, pharyngeal and esophageal phases. The timing of these phases is controlled by brain stem pattern generators including reflex for oropharyngeal propulsion and transit. Dysphagia in Parkinson’s disease (PD) commonly observed at late stages with aspiration, pneumonia and hospitalization.

**Objective:** Can subtle oromotor signs (if any) be observed for planning early interventions in PD

**Methods:** The present study investigated oromotor function in fourteen early PD (onset ≤2years; H&Y score ≤2.5) with dynamic MRI using saline (water) bolus and compared with seven age-matched healthy controls.

**Results:** All the patients with PD were non-symptomatic for dysphagia by self-reporting, and on clinical assessment (Part-II MDS-UPDRS, Swallowing disturbance questionnaire, SDQ and Clinical assessment of dysphagia in neurodegeneration, CADN). Qualitatively MR images visualized, differences in PD compared to healthy controls for tongue-wave, velar-closure or release, bolus placement, oropharyngeal reflex-initiation, transit-time, epiglottic-closure-coordination and post-swallow oral or pharyngeal residue. Descriptive analysis showed higher variability of velar-closure, oropharyngeal- and pharygoesophageal-transit time in patient with PD. Group analysis (two-sample) show significant difference for velar-closure.

**Conclusion:** Multiple lingual-waves, reverse-tongue thrust, with delayed velar control attributed to incoordinated muscular rhythm. Variable oropharyngeal transit time (0.64 to 2.25 msec) in PD ascribed to brainstem degenerative changes. Findings imply that subtle observable early oromotor signs as pre-clinical manifestation when evaluated with non-invasive, non-contrast dynamic MRI support early intervention, to prevent late-stage aspiration episodes and consequent hospitalizations.

## 1. Introduction

Swallowing, a basic oromotor function for survival may be affected in Parkinson’s Disease (PD) compromising the quality of life^1,2^. The prevalence of swallowing problems in neurodegenerative populations is under-reported as first episode of aspiration and pneumonia^3,4^. Prevalence of dysphagia in PD has been reported as 20% to 80%^3^ or 82% when the objective measures are used^5^. The causes of dysphagia in PD may be multifactorial, though muscle rigidity, slow, hesitant and incoordinated movements are major responsible factors^6^. These factors are difficult to evaluate as neither the occurrence nor severity of dysphagia correlate with overall disease^7,8^. Dietary habits, life-style may contribute to neurodegenerative progression^9,10^ and the oromotor function (speech) may play an important role in prodromal phase^11,12^. Early diagnosis of dysphagia in PD may prevent hospitalization, disability, deaths, and global burden^13^ for which periodical evaluation of oromotor function is required. Clinically early assessment is crucial for patient prognosis^14^ and available gold standard tools videofluoroscopy (VFS) or Flexible endoscopic evaluations of swallowing (FEES) are still with constraints ^2,15,16,17,18,19^. If such diagnosis is complemented with non-invasive, patient and environment friendly objective investigation, it will facilitate early identification, optimal intervention with better prognosis and reducing the morbidity.

Swallowing mechanism has four phases, problem may be in any phase or combination of phases in PD. The temporal tongue^20,21^, velopharyngeal^22^, pharyngeal wall and oesophageal impairments ascribed to ‘enteric nervous system’ are early modifications documented in PD^5^. With appropriate technique the subtle oromotor differences might be the first sign of the disease^5^ that go undiagnosed in early stages. Thus, present study was planned to investigate subtle oromotor signs during saline swallowing in early PD with non-invasive dynamic MRI.

## 2. Methods

After approval of the study from the Institute’s Ethics Committee, fourteen subjects (seven male and seven female) with Parkinson’s disease (PD) and seven healthy controls (four male and three female) of same age range were recruited from Neurology outpatient services. The inclusion criteria for group A (PD) subjects were: diagnosed with idiopathic PD by the neurologist and had given written informed consent for the investigations. The exclusion in group A were as-no other neurological disorder, psychiatric disorder, and contraindications for MRI. Inclusion criteria of group B (Healthy subjects, HC) were: no neurological, swallowing, psychiatric disorders, or any other disorder that can affect swallowing and had given written consent for the investigations. To rule out any other neurological disorder, MRI screening was also done for all the subjects. One of the healthy male control (HC) participants was excluded from the study after MRI due to extensive head motion.

### 2.1 Clinical Assessment

It included detailed patient’s history, Unified Parkinson’s Disease Rating Scale (MDS-UPDRS) and Hoehn and Yahr stage^23^ (H&Y; MDS, 2008). The subjects were elaborately interviewed for swallowing or any other oromotor problems (as self-report) scored on Swallowing Disturbance Questionnaire^4^ (SDQ), Clinical Assessment of Dysphagia in Neurodegeneration^24^ (CADN). SDQ is a fifteen item questionnaire regarding swallowing assessment in PD where more than eleven ‘yes’ has been considered as positive scores^4^. CADN is eleven item scale with two parts consisting of first as questionnaire (anamnesis - seven items) and second as clinical observation (four items) where more than one-and-half (>1.5) scores considered for further assessment. In degenerative diseases especially PD scoring of more than one-point-two-five (>1.25) is indicator for dysphagia investigation^24^. Disease signs and symptoms are clinically scored on MDS-UPDRS^23^ where Part-I evaluates ‘non-motor aspects of daily living experiences (nM-EDL)’, Part-II ‘motor aspects of daily living experiences (M-EDL)’, Part-III is the ‘motor examination’ by the investigator, and Part-IV assesses the ‘motor complications’.

### 2.2 Imaging technique

The MRI was performed on a 1.5T clinical MRI system (Magnetom Aera, Siemens Healthcare, Erlangen, Germany) using 12-channel head, 4-channel neck coil and large 4-channel flex body coils. General screening for any other neurological incidental finding was done with anatomical sequences including T1, T2, FLAIR and SWI/DWI. Subjects with incidental other neurological disorder were informed and excluded from the study.

Dynamic two-dimensional sagittal images were acquired using single shot T2 weighted trueFISP (fast imaging with steady state precision, trufi) sequence with cine-on and free breathing. The parameters for imaging were-field of view (FOV) 230, TR 162.54msec, TE 1.27msec, flip angle 44 degrees, matrix of 206 × 256, band width of 975, single slice with slice thickness of 10mm, acquisition 3.126 fps (frames per sec i.e. 0.32 sec per frame) and total acquisition time of 8.53 minutes (511.8 sec). The K-space acquisition was 80% with phase encoding direction as rows and 208 steps. As it was T2 weighted imaging, so normal saline appeared as bright contrast compared to soft tissues^25^.

#### Bolus

For swallowing the bolus was normal saline (NS, concentration 9 mg/ml i.e.0.9%) that was delivered through contrast media injector (Ulrich Medical, Ulrich GmbH & Co.KG, Germany), and sterile injector line where patient hose (250 cm) was placed in the oral cavity (fixed stably at 1cm inside, under the front incisors). Total NS was 71ml delivered in mouth with flow rate of 2ml/sec as - (i) first bolus of 21ml delivered in total injection time of 10.5 sec and later four bolus of 10 ml NS injection time of 5.0 sec. The interval after 21ml bolus was 120 sec and 65 sec inter-bolus interval for four 10 ml swallows to facilitate residual swallow efforts and post- swallow breathing.

### 2.3 Analysis of Data

Image processing of dynamic MRI was done in Syngo (Siemens provided) and ImageJ (Ferreira T, Rasb and W; https://imagej.nih.gov/ij) software. Transit time was calculated from movement completed in number of frames and the time of acquisition (frames per second). Analysis of clinical parameters was done with SPSS (version 23, IBM Corporation) software. Due to extensive head motion one of the male HC was excluded from the study analysis. Image rating was blinded during analysis and the images were qualitatively rated by two evaluators independently [blinded for images from group A (PD) or B (HC)].

## 3. Results

There were no significant demographic difference observed between two groups for age ranging 45 to 72 years (mean PD 60.43± 7.9 SD; HC 59.14± 8.3 SD) and gender (PD including 7M and 7F; HC including 4M and 3F). In patient group, mean duration of PD diagnosis was 1.75±1.5SD years and the disease was rated as mean 1.4±1.0SD H&Y stage (Table 1). On MDS-UPDRS rating mean motor function (Part-III) was observed as 21.22±1.50SD and 15.25±0.40SD as mean daily living motor experience (Part-II). Clinical questionnaire for swallowing (SDQ) was considered as positive when more than eleven items answered as ‘yes’^4^. None of the subject was observed as positive (that is scoring more than 11 yes). So, CADN^24^ was also used for clinical evaluation of swallowing where all the patients with PD scored ‘no’ (zero score), or ‘subclinical’ (one score) signs of dysphagia similar to the healthy control participants. Ten (71.43%) of PD subjects’ CADN score was ‘one’ and four (28.57%) of PD subjects’ CADN score was ‘zero’ whereas CADN was scored as ‘zero’ in 66.67% (four) healthy control subjects and 33.33% (two) [as ‘one’ of the HC was excluded from analysis]. None of the patients with PD scored CADN > 1.25 suggesting for further investigation.

**Table 1:** Clinical Scores Observed in PD patients (n = 14)

| Mean Disease Duration |  | MDS-UPDRS (Part II) |  | MDS-UPDRS (Part III) |  | H&Y |  |
| --- | --- | --- | --- | --- | --- | --- | --- |
| Mean | SD | Mean | SD | Mean | SD | Mean | SD |
| 1.75 | 1.5 | 15.25 | 0.40 | 21.22 | 1.50 | 1.81 | 0.92 |
| **Distribution of scores (percentage of individuals)** |  |  |  |  |  |  |  |
| H&Y |  | SDQ |  | CADN |  |  |  |
| ≤ 2 | >2 | < 11 scores | ≥11 scores | 0 score | 1 score | 2 score |  |
| 100 | 0 | 100 | 0 | 35.71 | 64.28 | 0 |  |

With dynamic MRI oromotor function was analysed for (i) pre-swallow articulatory posture, breathing pattern (pre- and post-swallow), (ii) air-swallow (dry), (iii) saliva-spooling, excessive-salivation, drooling and saliva-swallow and (iv) saline-swallow (Video). Dysphagia signs (water-like consistency) for two bolus volumes (21ml and 10ml) were rated separately to understand differences due to quantity^26^ and these signs were scored at four-point scale^27^ similar to modified barium swallow (MBS) (Table 1). The four 10ml boluses were rated and signs were considered as “dysphagia positive” when appeared on minimum 50% of swallow efforts with atleast mild score (Table 2). Comparing two groups with two-sample t-test the mean significant difference was found in velar closure (t = 3.41; p =0.004 two tailed) when equal variance not assumed (F = 16.64; p = 0.001) and mean difference for oropharyngeal transit time was observed but could not reach statistical significance (t=1.81; p = 0.08) equal variance assumed (F = 1.14; p = 0.299). Bolus volume splitting was similar in both HC and PD groups for 21ml swallow but 10ml bolus splitting was observed more frequent in PD (compared to HC).

**Table2:** Dysphagia Signs Observed in PD patients with dynamic MRI (n = 14; frequency of occurrence in percentage)

| S. No. | Abnormal Signs | No / Normal (0) | Mild (1) | Moderate (2) | Severe (3) |
| --- | --- | --- | --- | --- | --- |
| 1 | Labial closure | 85.71 | 14.28 | 0 | 0 |
| 2. | Tongue position | 28.57 | 7.14<br>(delayed)<br>50<br>(multiple efforts) | 14.28 | 0 |
| 3. | Bolus placement | 50 (7) | 50 (7) | 0 | 0 |
| 4. | Velopharyngeal closure | 42.85 (6) | 14.28 (2) | 42.86 (6) | 0 |
| 13. | Swallow reflex delay | 53.33 (7) | 33.28 (5) | 14.28 (2) | 0 |
| 14. | Oro-pharyngeal propulsion | 33.28 (5) | 53.33 (7) | 14.28 (2) | 0 |
|  | Oropharyngeal tiredness during last two 10ml boluses | 78.57 (11) | 21.43 (3) | 0 | 0 |
| 7. | Epiglottic movement | 78.57 (11) | 21.43 (3) | 0 | 0 |
|  | **Residue** | 33.28 (5) | 21.43 (3) | 33.28 (5) | 14.28 (2) |
| 15. | Posterior tongue |  | 21.43 (3) |  |  |
| 16. | Oropharangeal |  |  | 33.28 (5) |  |
| 17. | Esophageal |  |  |  | 14.28 (2) |
|  | **Post-swallow Clearance** |  |  |  |  |
| 18. | Residue / Dry / saliva | 33.28 (5) | 21.43 (3) | 38.15(5) | 7.14 (1) |
| 19. | Residue clearing efforts | 42.86 | 28.43 | 14.28 | 14.28 |
|  | **Penetration** |  |  |  |  |
| 20. | Premature loss of bolus | 85.71 | 7.14 | 7.14 | 0 |
| 21. | Nasal regurgitation | 100 | 0 | 0 | 0 |
| 22. | Laryngeal aspiration | 100 | 0 | 0 | 0 |
|  | **Other observations** |  |  |  |  |
| 23. | Coordination of velopharyngeal-laryngeal movements | 7.14 | 64.28 | 28.43 | 0 |
| 24. | Pre-swallow Saliva-spooling in oral cavity | 85.71 | 14.28 | 0 | 0 |
| 25. | Excessive Saliva (sialorrhea or drooling) | 100 | 0 | 0 | 0 |
| 25. | Pre-swallow articulatory posture | 78.57 | 14.28 | 7.14 | 0 |
| 26. | Velum-Epiglottic-Laryngeal posture during breathing | 92.86 | 7.14 | 0 | 0 |

Scoring was done as four-point scale with zero (0) as - no abnormality / Normal, one (1)- Mildly affected, two (2)- Moderately affected, and three (3) as Severely affected. [*\*Inspired by VFS Worksheet for Swallowing (VEWS) Video Swallow/Modified Barium Swallow (MBS) VA Boston Healthcare (Gramigna, 2006)*]

Descriptive analysis showed differences in oral, oropharyngeal and pharyngeal phases in patients with PD (Table 3). In PD group the oropharyngeal transit time ranged from 0.32 msec to 1.28 msec while HC the variation was only of one msec. Similar range variations were also observed for phayngoesophageal and esophageal transit time, though the group-means were non-significantly different. Duration of tongue wave (lingual wave) for bolus movement in oral phase varied in HC group higher as compared to PD (range HC = 1.47, PD = 1.00).

**Table 3:**
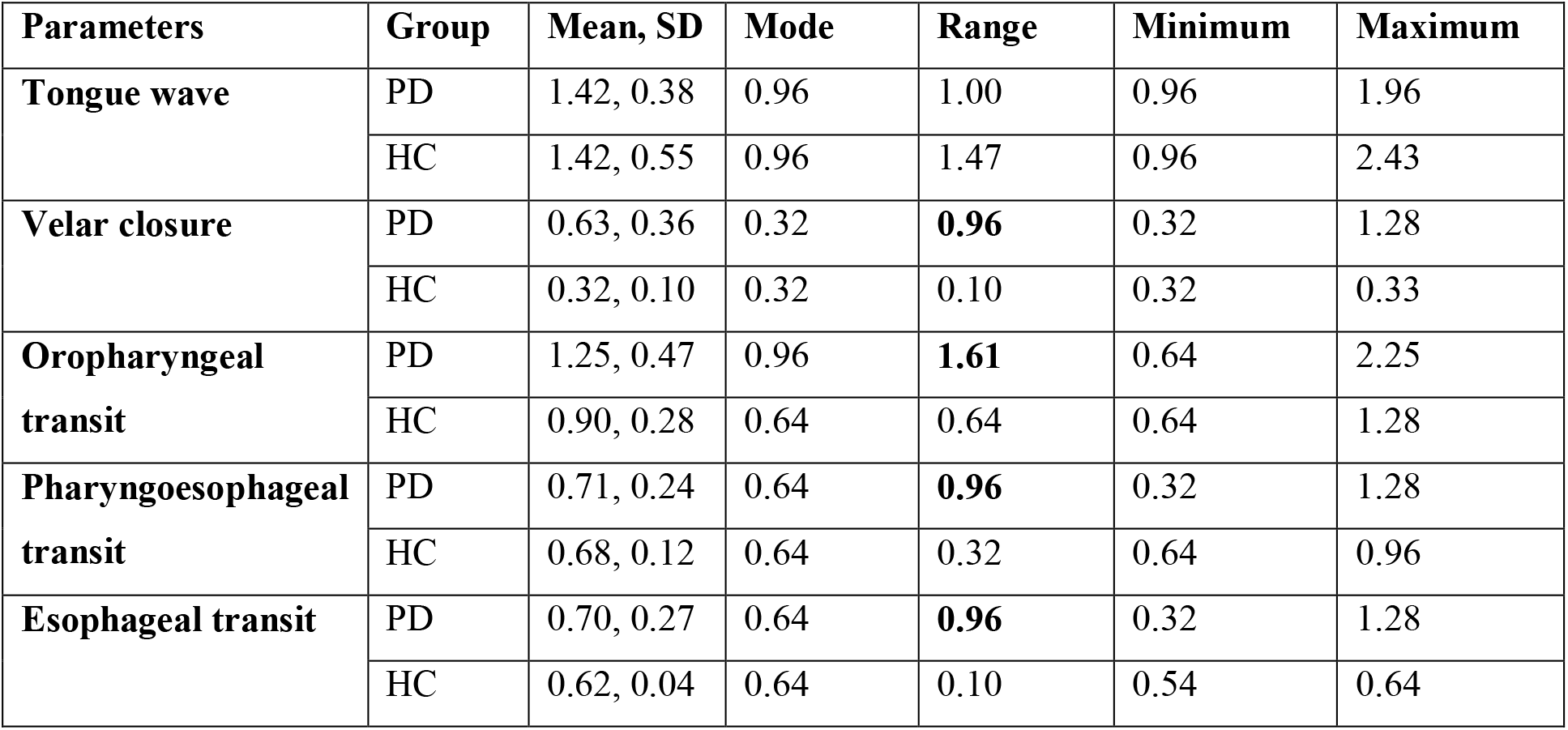
Descriptive findings of dynamic MRI parameters (PD n = 14; HC = 6)

Qualitatively early tongue positioning, multiple lingual-wave efforts, delay in posterior-push, and post-swallow residue was observed in individuals with PD during oral phase (Table 2). In oropharyngeal phase slow velar-elevation or release, incomplete-velar-closure, swallow-response-delay (reflex), delayed transit time, post-swallow-residue and multiple residue-swallow-efforts were noted. One of the PD patients had multiple oropharyngeal-propulsion efforts. Nasal regurgitation was not observed in any of the subjects (neither HC nor PD).

Pharyngeal phase involved differences in PD patients for posterior wall peristalsis, transit-time, tongue-root hyoid-bone coordination, epiglottic-closure, laryngeal-closure, post-swallow residue and residue-swallow-efforts. Resuming post-swallow breathing coordination during oesophageal bolus transit was visible with epiglottic-arytenoid-movement or opening (Figure 1, A). It was observed that two of the PD patients had ‘bolus residue stuck on epiglottic dorsal wall’ during post-swallow breathing (when epiglottis and arytenoids were open) for multiple 10ml boluses (Figure 1, B).

**Figure 1(A).**
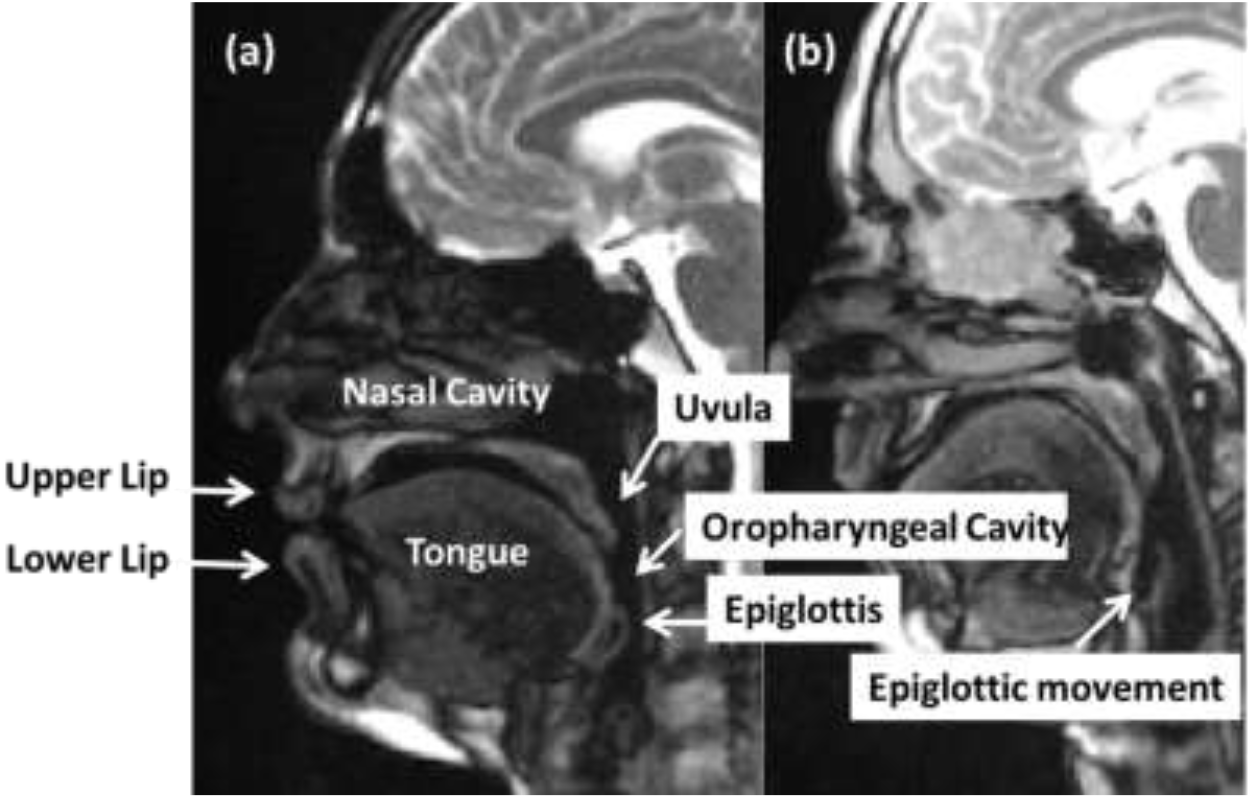
Anatomical structures visualized with dynamic MRI during swallowing and breathing.

**Figure 1(B).**
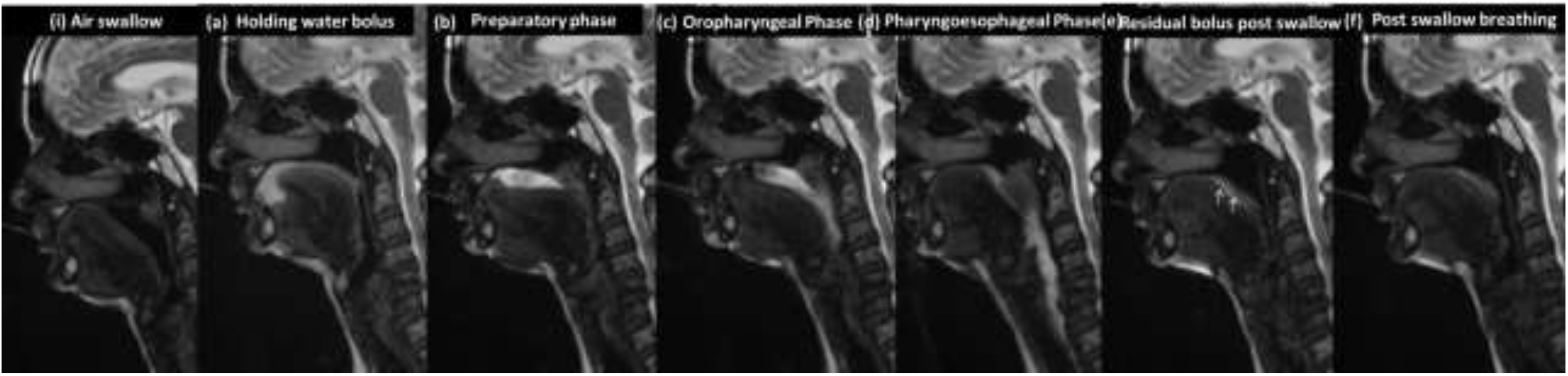
Swallowing phases and signs of dysphagia in subject with Parkinson’s disease (2D slices from dynamic MRI)

## 4. Discussion

The aim of the study was to evaluate presence of (if any) subtle oromotor signs that may support early intervention and prevent late stage dysphagia complications in PD. During this study MRI dynamic imaging was done using saline bolus (water like consistency) with two volume sizes to unfold the swallowing mechanism in patient diagnosed with PD, when clinically there were no significant signs of dysphagia. In present study, early signs of dysphagia in PD were elaborated with T2 weighted dynamic MR images visualising soft tissue contrast for tongue movement (tip, dorsum and base), velopharyngeal sphincter action, pharyngeal propulsion, epiglottic inversion, laryngeal elevation, pharyngeal peristalsis and respiratory coordination^28^. Findings of slow-velar closure, delayed backward propulsion, multiple lingual-efforts in PD suggested hesitation of motor-execution or motor rigidity^29^ when corroborated to oropharyngeal coordination (delayed transit-time, post-swallow residue, multiple residue-swallow-efforts) may be attributed to locus coeruleus (brainstem) and/or overall sensorimotor control (including limb-motor)^30,31^.

Bolus splitting differences in PD and healthy control for 21ml and 10 ml volume is in concordant to previous studies though involving different techniques^13,32^. In present study mean group comparison does not show statistical difference for lingual wave (oral phase) though multiple efforts to position the bolus for backward propulsion indicated motor hesitation, rigidity and bradykinesia^6^. Reverse (forward) tongue thrust was observed in three PD patients suggested altered oromotor control in asymptomatic dysphagia stage^30,31^.

Post-swallow oral residue in posterior oral cavity may be attributed to tongue-rigidity, inadequate pressure against palate and sensorimotor control^33^. These difficulties with bolus control in oral phase and premature loss to the pharynx in early PD are attributed to locus coeruleus^30^ or nor-adrenergic mechanisms in brainstem^30,31^. Delayed velar raising and inadequate velar closure before initiation of backward propulsion showed incoordinated muscular rhythm in six patients. In PD the tongue function in bolus formation have been reported to be associated with oropharyngeal transit time^31,34^. In present study oropharyngeal transit time (as defined Namasivayam-MacDonald)^35^ was increased in many patients with PD though mean group comparison could not reach significant difference. It may be clarified with large variation of transit-time in PD group (0.64 to 2.25 msec). Swallow response (reflex) delay in oropharyngeal phase and increased transit time has been ascribed to brainstem degenerative changes^36,37,38^. It may also be the altered sensory feedback modifying the brainstem reflexes of oropharynx and oesophagus^38,40^. Delayed bolus transport to pharynx with altered pharyngeal peristaltic movement may be excitatory-inhibitory reflex rhythm modification^38^ as early marker in PD^40,41^. These brainstem reflex rhythms might improve with cervical electric stimulation therapies^42^.

Differential diagnosis of swallowing phases and subtle associated signs in preclinical stage may support tailoring treatment strategies in PD (Figure 1, B). Differentiation oromotor signs in clinical stages may also facilitate optimizing the dopamine replacement therapies^43^ and planning the site of deep brain stimulation^44^. Muscle-strengthening, movement-range and sensory-perceptual exercises improve lingual-wave delay and/or bolus control in oral phase^5,45^ due central-peripheral mechanisms instead of muscular changes^46^. Intensive voice treatment (LSVT) targeting tongue root movement may improve coordination between breathing and swallowing or airway protection^47^. Customizing the management depending on visualized deficit signs will facilitate optimal management for example (i)patients with velar-closure delay/incomplete, if intervened with velopharyngeal sensory stimulation or muscle strength exercises; (ii)patients with oropharyngeal transit delay may be guided with muscle-coordination and perceptiomotor reflex stimulation and (iii)in cases of incoordinated epiglottic opening, voluntary cough (expiratory muscle strength training) may help to eject and decrease chance of penetration / aspiration^5^.

Incoordinated breathing pattern, hesitant motor initiation, inadequate oropharyngeal clearance and pharyngeal residue were potential aspiration risks observed in the study which may have remained underdiagnosed^48^. These signs were unnoticed by themselves (patients) and could not be assessed with available standardized clinical examination (SDQ and CADN). MDS-UPDRS part II (Motor experiences of daily living) and MDS-UPDRS part III (Motor examination) mean scores of PD group (Table 1) disclose mild stage of disease^49^. It was discordant with oromotor signs, demonstrating the preclinical empirical-evidence for diagnosis and intervention. It necessitates incorporating detailed examination protocols for early detection and prevention of dysphagia or aspiration in PD. MDS-UPDRS part III examines overall severity, suggesting the impairment of cortico-striatal loops as key phenomenon for motor symptoms in PD^31,50^ where oromotor subtle information may characterize to such changes^30^ that are non-differentiable in present tool, further designate that all the patient with PD even in early stages, need the extensive assessment of these functions.

Future animal studies and human studies with bigger sample, may facilitate correlation of oromotor function with neural-degeneration. Expanding differences in boluses consistency and texture during swallowing with thickened purees (semisolids with varying density) and mastication pattern with water-rich fruits may further unfold asymptomatic stage of dysphagia in early PD. Dynamic studies for the physiological differences, tolerance, residue and aspiration due to increased/decreased texture-cohesion or surface-tension may support variation assessment in the cohort. Laryngotracheopulmonary aspiration (aryepiglottic, ventricular and vocal folds closure)^51^ may further be elaborated revealing airway protection and risks.

### Advantages

All the patient with PD were asymptomatic clinically and reported no problem in swallowing still many of them were at risk of dysphagia and aspiration, the preclinical and prediagnostic phase^48^. Visualization of subtle oromotor signs (with dMRI) which were not accounted with clinical test (SDQ, CADN, UPDRS-MDS), facilitate the early intervention. The study enabled standardizing the non-invasive tool (dynamic MRI) that overcomes the limitations of clinical gold standard technique of VFS with barium^27^, patient specific medical contraindication for radioactive drug and radiation exposure (limiting evaluation sessions). As compared to FEES studies it benefits to assess oral and oropharyngeal signs in early PD. It explicitly discriminated all the phases and qualitatively elaborate signs during swallowing for customized appropriate optimal management. In future it may facilitate the differential diagnosis of Parkinson plus syndrome in early stages^51,52^.

### Limitations

It was pilot study with small sample (though with sound methodology), resulting in few limitation of the study. The study could not differentiate patient subgroups with PD stages (H&Y 1, 1.5, 2 and 2.5) or patients with and without dysphagia risk-signs. Dynamic MRI constraints in swallowing assessment are: (a) supine position in some cases, it may be awkward or unnatural position for water-intake (b) it may not fully represent actual situation, though it has been reported as noninterfering with assessment^28,53,54^; (c) may not be feasible in medically unstable, disoriented, or uncooperative patients like VFS and FEES; (d) objectifying the overlapping phases of swallowing; (e) patients with MRI contraindications cannot be evaluated.

## 5. Conclusion

Findings show that PD patients in early stages (<2.5 H&Y) are clinically asymptomatic (the prediagnostic phase) where subtle oromotor signs are detected with dynamic MRI, a non-invasive tool (without any irradiation). The uncoordinated airway dynamics before, during, and after swallowing envisage the risk of dysphagia and aspiration. Objective early diagnosis will facilitate the customized optimal remediation, preventing life threatening events and may in future unfold prodromal oromotor signs of airway, aspiration and dysphagia in PD.

## Data Availability

The data may be shared on appropriate email request

